# Discovery of SARS-CoV-2 strain of P.1 lineage harboring K417T/ E484K / N501Y by whole genome sequencing in the city, Japan

**DOI:** 10.1101/2021.02.24.21251892

**Authors:** Yosuke Hirotsu, Masao Omata

## Abstract

On the February 2020, the very first case was an American female from Diamond Princess cruise ship. Since, we have confirmed 136 patients infected with coronavirus disease 2019 (COVID-19) until February 2021. Here, we conducted the whole genome sequencing analysis of severe acute respiratory syndrome coronavirus 2 (SARS-CoV-2) on samples from 70 of 136 patients (51.5%). These patients were infected in Diamond Princess cruise ship (n=1), Africa (n=2), Japan (n=66) and Brazil (n=1). The viral genome sequence of a patient on the Diamond Princess cruise ship in February 2020 was similar to that of original strain found in Wuhan, China (19A clade). Four patients, including two returnees from Africa and two lived in Japan, confirmed at the end of March 2020 had sequences similar to those of lineage with D614G mutation, which was endemic in Europe (20A [n=3] and 20B [n=1] clade). The 64 Japanese patients confirmed from September 2020 to January 2021 had sequences similar to those of the currently prevalent lineage (20B [n=58] and 20C clade [n=6]). Subsequent analysis revealed three mutations (K417T/ E484K / N501Y) in the receptor binding domain of the spike protein in a man in his 40s. The sequence was identical to the P.1 lineage (also known as 20J/501Y.V3) reported in Brazil. This is the first report of SARS-CoV-2 P.1 lineage identified in the city, Japan.

## Introduction

Severe acute respiratory syndrome coronavirus 2 (SARS-CoV-2) was first identified in Wuhan province, China, at December 2019. The virus is highly transmissible and its basis reproduction number (R_0_) is estimated to range between 2.2 and 3.9 [1]. To date, 107 million individuals have been infected with SARS-CoV-2 and 2.4 million patients have passed away from coronavirus disease 2019 (COVID-19) [2].

SARS-CoV-2 belong to the *Coronaviridae* family and subdivided into genera β-coronaviruses, subgenus *Sarbecovirus*. SARS-CoV-2 has a single-stranded, positive-sense RNA genome with a total of 29.9 kilo base pairs in length. SARS-CoV-2 has the largest genome size among RNA virus and encodes for a 3′-5′ exoribonuclease with proofreading function, which result in low mutation rate compared to other RNA virus [3].

It is assumed that most of the mutations occur neutrally and may not influence on virus properties. However, pandemic spread of the virus in naïve populations may affect on the selection of mutations that alter pathogenesis, virulence, and/or transmissibility [4, 5]. Currently, an emergent D614G mutation in the spike glycoprotein of SARS-CoV-2 is prevalent globally [6, 7]. Afterward, new emerging lineages with spike protein mutations were discovered in United Kingdom (B.1.1.7 lineage, 20I/501Y.V1, also named VOC 202012/01) [8], South Africa (B.1.351 lineage, 20H/501Y.V2) [9] and Brazil (P.1 lineage, 20J/501Y.V3) [10, 11]. All of these lineages have N501Y mutation in receptor binding domain (RBD), which directly binds to the angiotensin converting enzyme 2 (ACE2) receptor on host cell [12]. N501Y mutation would contribute to the high transmissibility [13, 14]. The B.1.351 and P.1 lineages have K417N/T and E484K mutations along with N501Y in RBD of spike protein.

In this study, we performed the whole genome sequencing using 70 nasopharyngeal swab samples from COVID-19 patient collected between February 2020 and February 2021. Of 70 samples, we identified SARS-CoV-2 P.1 lineage in the Kofu city, Japan.

## Methods

### Patients and sample collection

A total of 136 COVID-19 patients were confirmed in our hospital. Of these, we included 70 samples for subsequent genome analysis. For these patients, one was a passenger on a Diamond cruise ship on February 2020, four was confirmed at our district on the end of March 2020, 64 was confirmed on September 2020 to January 2021, and one was on February 2020. These included one American patient infected in Diamond Princess cruise ship, two returnees from Africa, one returnee from Brazil and 66 Japanese, one year past from first confirmed case.

Nasopharyngeal swab samples were collected using cotton swabs and placed in 3 mL of viral transport media (VTM) purchased from Copan Diagnostics (Murrieta, CA, USA). We used 200 μL of the VTM were used for nucleic acid extraction within 2 hours after sample collection.

### Viral nucleic acid extraction

Total nucleic acid was isolated from the samples using the MagMAX Viral/Pathogen Nucleic Acid Isolation Kit (Thermo Fisher Scientific, Waltham, MA, USA) on the KingFisher Duo Prime System (Thermo Fisher Scientific) as we previously described [15, 16]. Briefly, we added 200 µL of VTM, 5 µL of proteinase K, 265 μL of binding solution, 10 μL of total nucleic acid-binding beads, 0.5 mL of wash buffer, and 0.5–1 mL of 80% ethanol to each well of a deep-well 96-well plate. The nucleic acids were eluted with 70 μL of elution buffer.

### Quantitative reverse transcription PCR (RT-qPCR)

According to the protocol developed by the National Institute of Infectious Diseases (NIID) in Japan [15, 17-19], we performed one-step RT-qPCR to detect SARS-CoV-2. This PCR amplifies the *nucleocapsid* (*N*) gene of SARS-CoV-2 (NC_045512.2). For the internal positive control, the human ribonuclease P protein subunit p30 (*RPP30*) gene was used (Integrated DNA Technologies, Coralville, IA, USA) [3]. The RT-qPCR assays were conducted on a StepOnePlus Real-Time PCR System (Thermo Fisher Scientific) with the following cycling conditions: 50°C for 5 min for reverse transcription, 95°C for 20 s, and 45 cycles of 95°C for 3 s and 60°C for 30 s. The absolute copy number of viral loads was determined using serial diluted DNA control targeting the *N* gene of SARS-CoV-2 (Integrated DNA Technologies) as previously described [18].

### Antigen test

The sample antigen levels were determined quantitatively with the LUMIPULSE SARS-CoV-2 Ag test (Fujirebio, Inc., Tokyo, Japan) according to the manufacturer’s instructions [20-22]. In brief, 700 μL of the VTM samples were briefly vortexed, transferred into a sterile tube, and centrifuged at 2,000 ×*g* for 5 min. Aliquots (100 μL) of the supernatant were used for testing on the LUMIPULSE G600II automated system (Fujirebio). For samples with an antigen level > 5,000 pg/mL, the samples were diluted with the kit diluent and re-tested, and the antigen level was calculated taking the dilution factor into account. Samples with an antigen level ≥ 10 pg/mL were considered positive, samples with ≥ 1.0 pg/mL and < 10.0 pg/mL antigen were labeled inconclusive, while a result of <1.0 pg/mL was considered negative as per the manufacturer’s guidelines [22].

### Whole genome sequencing

We amplified the genomic region of SARS-CoV-2 using the Ion AmpliSeq SARS-CoV-2 Research Panel consists of two primer pools covering 99.7% of the viral genome (Thermo Fisher Scientific). The amplicon size ranges from 125 to 275 base pairs in length. Extracted nucleic acids were subjected to genome sequencing on the Ion Torrent Genexus System, which automates the specimen-to-report workflow and yields results in a single day.

### Sequencing data analysis

The data of sequencing reads and quality control were processed on Genexus Software with SARS-CoV-2 plugins. The sequencing reads were mapped and aligned on the reference genome of SARS-CoV-2 strain Wuhan-Hu-1 (Accession no NC_045512) using the torrent mapping alignment program (TMAP). After initial mapping, a variant call was performed using the Torrent Variant Caller. The COVID19AnnotateSnpEff plugin was used for annotation of variants. Assembly was performed with the Iterative Refinement Meta-Assembler (IRMA) [23], which produced the FASTA file.

### Phylogenic tree analyses and lineage classification

Phylogenetic tree analysis and classification were conducted using Nextstrain [24] and Global Initiative on Sharing Avian Influenza Data (GISAID) database [25]. The Nextclade beta (https://clades.nextstrain.org/) was used to classify and generate phylogeny of virus strain. ‘hCoV-19/Wuhan/WIV04/2019’ strain isolated from Wuhan, China used as reference for detecting variants [26].

FASTA files of SARS-CoV-2 sequence were uploaded on the Nextclade. The nomenclature used by Nextstrain to designate clades are defined by specific signature mutations (https://nextstrain.org/blog/2021-01-06-updated-SARS-CoV-2-clade-naming). The international clade nomenclature system was used according to the Nextstrain.org (https://virological.org/t/updated-nextstain-sars-cov-2-clade-naming-strategy/581). The viral lineages was referred as Phylogenetic Assignment of Named Global Outbreak Lineages (Pangolin; https://cov-lineages.org/index.html) [27].The consensus genome sequence of identified P.1 lineage strain was deposited to the GISAID EpiCoV database with accession (EPI_ISL_978917).

Global sequencing data of P.1 lineage was exported from GISAID EpiCoV database by 14^th^ February, 2021. We searched and found the 121 available datasets. We used these metadata and FASTA files were used for subsequent analysis.

### Ethical statement

The Institutional Review Board of the Clinical Research and Genome Research Committee at Yamanashi Central Hospital approved this study and the use of an opt-out consent method (Approval No. C2019-30). The requirement for written informed consent was waived owing to the observational study and the urgent need to collect data of COVID-19. Participation in the study by patients was optional. All methods were performed in accordance with the relevant guidelines and regulations, and with the Helsinki Declaration.

## Results

### Landscape of genomic characterization and phylogeny

To determine the genomic characterization of SARS-CoV-2 identified in the Kofu city, Japan, we started to whole genome sequencing analysis since 8^th^ January, 2021. A total of the 136 samples obtained from COVID-19 patients who were confirmed in our hospital. Of these, 70 samples were subjected to analysis, which represented 51.5% of the total confirmed patients by February 15^th^ 2021.

The yielded sequence data was subjected to phylogenetic analysis using the Nextclade. As a result, we identified five types of clades including 19A clade (n=1), 20A (n=3), 20B (n=59), 20C (n=6) and 20J/501Y.V3 (n=1) (Figure 1).

**Figure 1.**
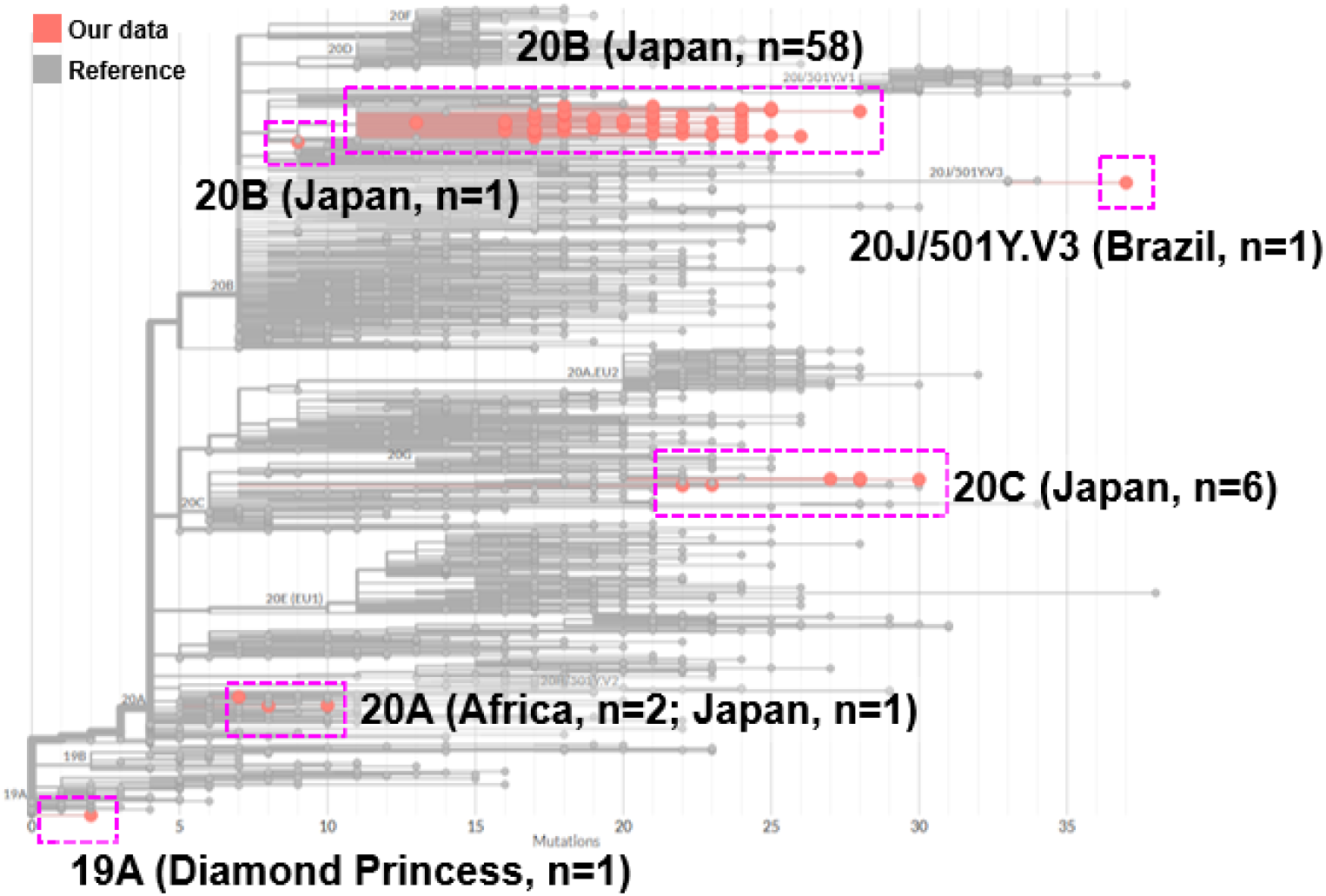
Phylogenetic tree analysis of SARS-CoV-2. Sequencing data was uploaded to the Nextclade (https://clades.nextstrain.org/) and visualized onto the location of phylogeny. The pink dotted line shows the sequencing data obtained in this analysis.

The SARS-CoV-2 from a patient on Diamond cruise ship was classified into 19A clade (Figure 2), and four patients confirmed on the end of March 2020 were classified into 20A (n=3) and 20B (n=1). From September 2020 to January 2021, SARS-CoV-2 from 64 patients were classified into 20B clade (n=58) and 20C (n=6) (Figure 2). Newly confirmed patient was classified as 20J/501Y.V3 (P.1 lineage).

**Figure 2.**
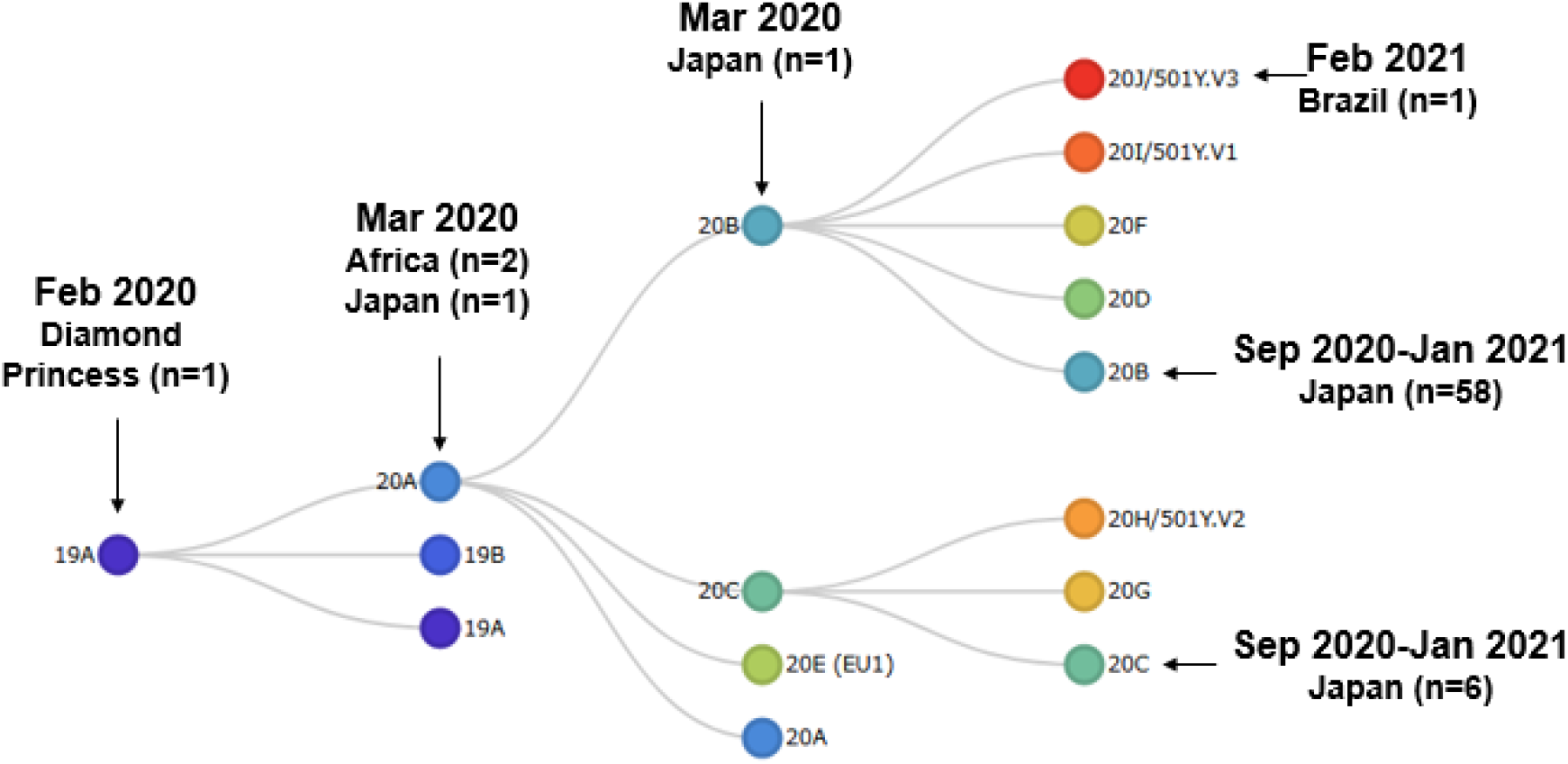
The relationship between the SARS-CoV-2 clade and timeline. The schematic tree shows of the Nextstrain clade in website. The arrows indicate the clade into which the 70 patients identified in this analysis were classified.

### Identify a patient infected with SARS-CoV-2 of 20J/501Y.V3 (P.1 lineage)

A man in his 40s, who had a history of staying in Brazil, attended our hospital in early February with symptoms of fever at 38.9 °C. RT-qPCR judged positive with high viral load (7.1 log_10_/µL) and low Ct value (Ct: 15). The antigen level was also high at 183,426 pg/mL. The patient was confirmed to be negative for SARS-CoV-2 by airport quarantine when he returned to Japan four days earlier. His symptom was mild and he was admitted to another hospital.

Sequencing analysis revealed that the SARS-CoV-2 of 20J/501Y.V3 (P.1 lineage) had 37 mutations including 22 missense, 10 synonymous, 3 intergenic, one frameshift and one in-frameshift mutation. In spike protein, we observed 12 missense mutations (L18F, T20N, P26S, D138Y, R190S, K417T, E484K, N501Y, D614G, H655Y, T1027I and V1176F).

These mutations were perfectly matched with the mutations in P.1 lineage previously discovered in Brazil [10] (Figure 3). In RBD of spike protein, three mutations (K417T, E484K and N501Y) were identified. These results suggested we identified the emerging strain related to 20J/501Y.V3 (P.1 lineage) in Japan.

**Figure 3.**
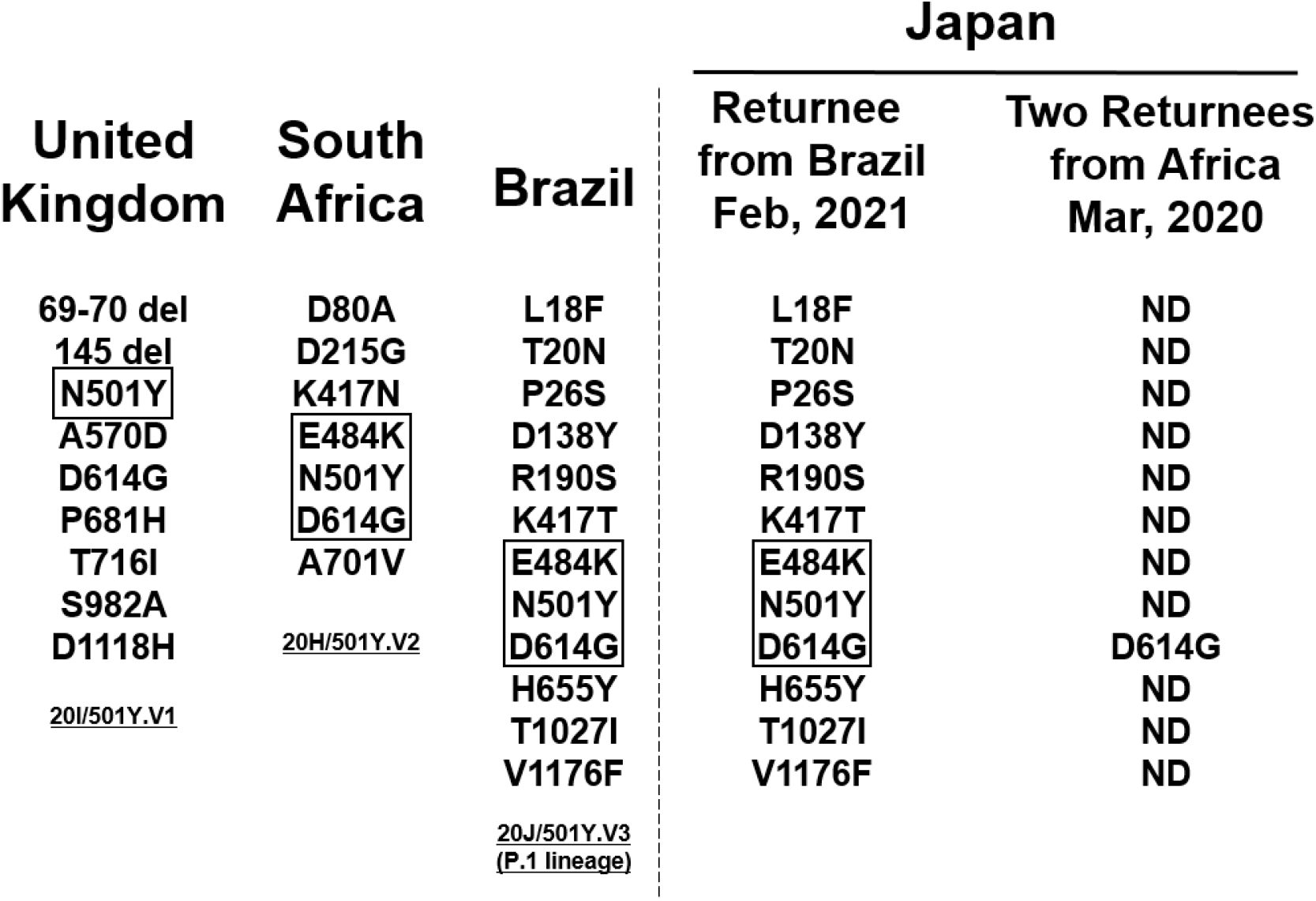
Mutations in SARS-CoV-2 spike protein. The left side of the dotted line shows the amino acid changes identified in the emerging strains reported in the United Kingdom, South Africa, and Brazil. The right of the dotted line shows the results of the current analysis: the patient who returned from Brazil on February, 2021 had the same mutation as 20J/501Y.V3 (P.1 lineage); the two patients who returned from Africa on March, 2020 had only D614G mutation in spike protein. The highlighted areas surrounded by lines indicate mutations in the receptor binding domain. ND, not detected.

### Global genomic surveillance of 20J/501Y.V3 (P.1 lineage)

To examine the global data of SARS-CoV-2 of 20J/501Y.V3 (P.1 lineage), we referred the sequence data deposited in GISAID [25, 28]. By February 14^th^ 2021, a total of 121 sequence data was available, and 119 were derived from patient and 2 were viral strains isolated from TMPRSS2-expressing Vero E6 cell line [29].

The SARS-CoV-2 of 20J/501Y.V3 (P.1 lineage) was first discovered in the sample collected December 4^th^, 2020 Manaus, Amazonas state in Brazil (Figure 4A). Afterward, the strain of 20J/501Y.V3 (P.1 lineage) have been continuously identified over the time (Figure 4A).

**Figure 4.**
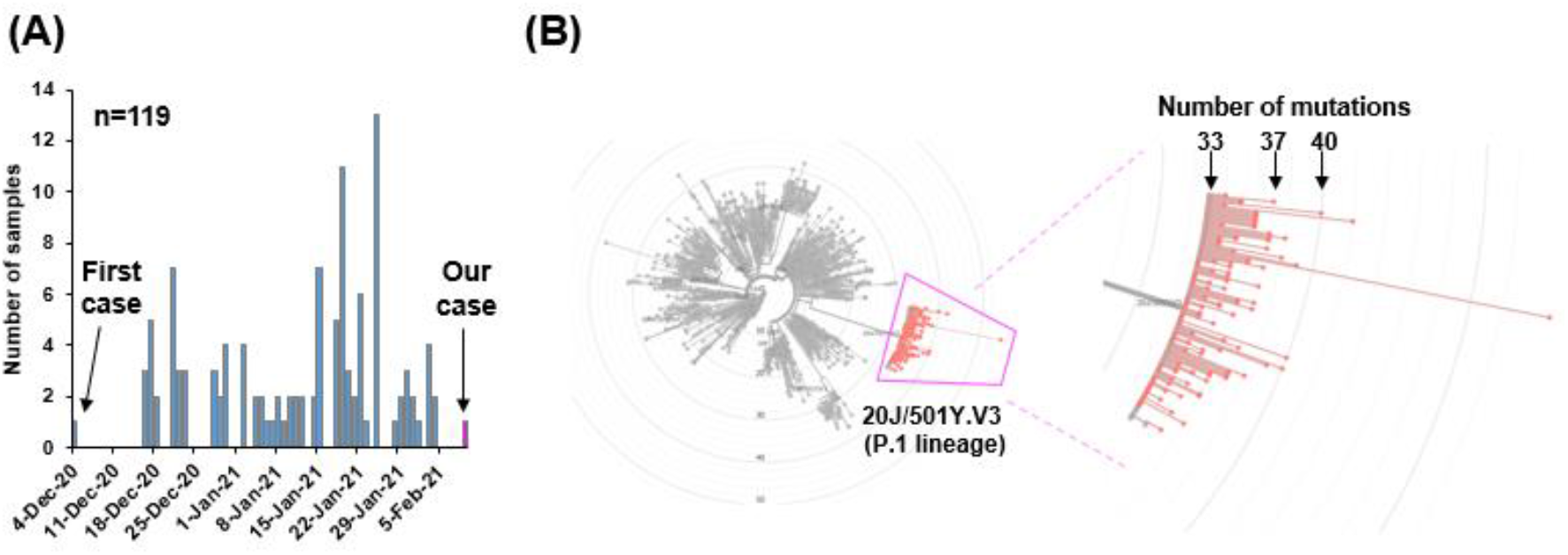
Global data of 20J/501Y.V3 (P.1 lineage) deposited in GASAID. **(A)** The number of samples of 20J/501Y.V3 (P.1 lineage) strain deposited in GISAID by February 14^th^ 2021. The first case was identified in December 4^th^, 2020. **(B)** A total of 119 sequencing data were analyzed on Nextclade. The radial phylogenetic tree shows the location of 20J/501Y.V3 (P.1 lineage) (left diagram). The magnified version of image shows the P.1 lineage surrounded by pink lines (right diagram). The total number of mutation was denoted when the SARS-CoV-2 strain Wuhan, China used as a reference.

Almost of 20J/501Y.V3 (P.1 lineage) have 33-40 mutations compared with original strain reported from Wuhan, China [26], and our identified strain had 37 mutations (Figure 4B). Of 119 patients, 82 (68.9%) were identified in Brazil, 5 (4.2%) in Japan, 20 (16.8%) in Europe, 3 (2.5%) in USA (Table 1), suggesting P.1 lineage began to spread in the world.

**Table 1.**
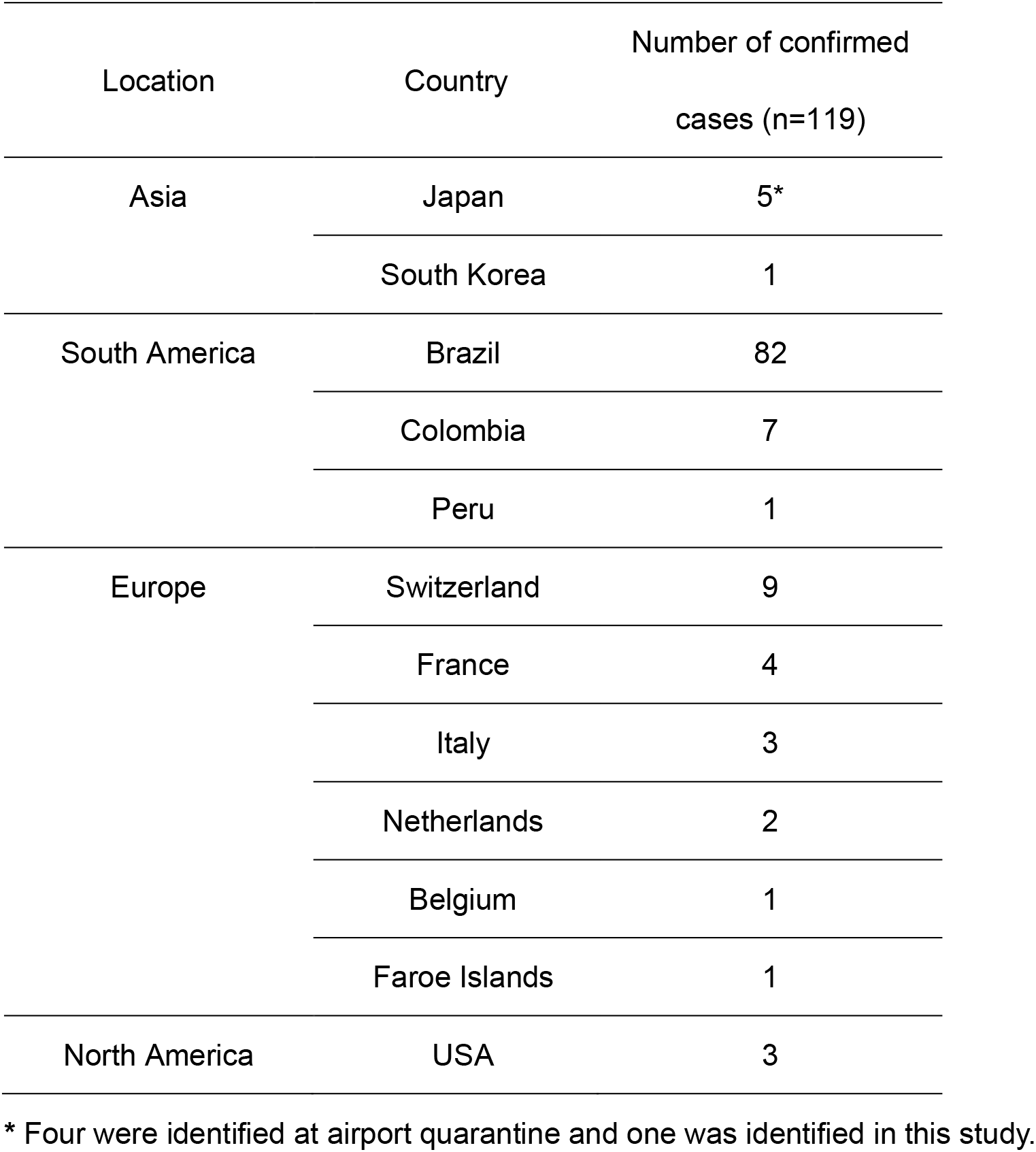
The prevalence of SARS-CoV-2 of 20J/501Y.V3 (P.1 lineage) in the world.

## Discussion

We started to the genomic surveillance to monitor the SARS-CoV-2 variants from 8^th^January, 2021. In this study, we confirmed that five clades have emerged over the time. The consecutive analysis identified SARS-CoV-2 of 20J/501Y.V3 (P.1 lineage) in a patient and detected mutations were identical to those of original P.1 lineage discovered in Brazil [10]. This is the first report on the 20J/501Y.V3 (P.1 lineage) in the city, Japan.

On January 6, 2021, the NIID detected a new emerging strain (P.1 lineage) of SARS-CoV-2 at airport quarantine in four travelers who arrived from Brazil on 2^nd^ January 2021 [30-32] (Table 1). Subsequently, almost of P.1 lineage discovered in Manaus in Brazil and gradually spread worldwide [10, 11]. This lineage has several mutations in spike protein and some of these mutations are shared with as 20I/501Y.V1 (B.1.1.7 lineage) in Unites Kingdom and 501Y.V2 (B.1.351 lineage) in South Africa.

In particular, mutations in the RBD of spike protein are noteworthy [33, 34]. N501Y mutation increases the overall binding affinity between RBD and human ACE2 receptor [35, 36]. Recently, escape mutations from neutralizing antibody recognition were identified by SARS*-*CoV-2 spike protein-expression vesicular stomatitis virus (VSV) and yeast [37, 38]. The K417N/T, E484K and N501Y mutation reduced the neutralizing activity of convalescent and mRNA vaccine-elicited serum [39-43].

The SARS-CoV-2 variant is not always related to threat to human health because the virus acquires genomic diversity during the course of its life cycle [44]. However, some of these mutations would be associated to attenuate the neutralizing activity of antibody. During the ongoing evolution of SARS-CoV-2, the new emerging lineage is likely to be circulating in the human population. Thus, genomic surveillance is important for public health to monitor the emerging lineage, evaluate vaccine efficacy and virus transmissibility.

## Data Availability

Data is available upon request.

## Acknowledgments

This study was supported by a Grant-in-Aid for the Genome Research Project from Yamanashi Prefecture (to M.O. and Y.H.), the Japan Society for the Promotion of Science (JSPS) KAKENHI Early-Career Scientists JP18K16292 (to Y.H.), a Grant-in-Aid for Scientific Research (B) 20H03668 (to Y.H.), a Research Grant for Young Scholars (to Y.H.), the YASUDA Medical Foundation (to Y.H.), the Uehara Memorial Foundation (to Y.H.), and Medical Research Grants from the Takeda Science Foundation (to Y.H.). We thank Hitoshi Mochizuki for applying for the ethics committee. We also thank Masato Kondo, Ryota Tanaka and Kazuo Sakai (Thermo Fisher Scientific) for technical help and all of the medical and ancillary hospital staff and the patients for consenting to participate.

